# Estimation of the Variation in Glomerular Filtration Rate Based on Glycosylated Hemoglobin, Serum Creatinine and Age, in Type 2 Diabetic Patients with or Without Chronic Kidney Disease

**DOI:** 10.1101/2023.02.17.22276261

**Authors:** Wilfor Aguirre-Quispe, Cesar Alejandro Arana-Calderón

## Abstract

**Objective:** The methods currently used to calculate GFR underestimate this measurement in the population of diabetic patients, so there is a need to look for more accurate methods of estimating GFR in this specific population. This study aims to evaluate a predictive model based on the use of HbA1c to estimate the variability of GFR in diabetic patients with or without CKD.

**Methods:** We analyzed data from diabetic patients belonging to a cohort of prospective follow-up of a renal health surveillance program attached to a Peruvian hospital. The following factors were included in the multiple linear regression model: age, sex, diastolic blood pressure (DBP), systolic blood pressure (SBP), body mass index (BMI), cholesterol, triglycerides, HDL, LDL, serum creatinine, Urinary creatinine, microalbuminuria, hemoglobin, basal glycemia and HbA1c.

**Results:** 122 patients were included in the analysis. The final multivariate model, which included variation of HbA1c, age and creatinine variation, was very significant (p <0.0001) with an adjusted R2 of 80%. The other variables analyzed were not significant to predict the variation of the GFR despite showing some correlation.

**Conclusions:** The study shows that HbA1c, age and creatinine variation significantly predict the variation of GFR in diabetic patients with or without CKD and opens the possibility of use as a prognostic tool for this specific population.

## INTRODUCTION

Chronic kidney disease (CKD) affects about 50% of the global diabetes population [1], with rates ranging from 40% to 83% in developed countries [2-4]. It is estimated that the prevalence of CKD in patients with diabetes will continue to increase dramatically over the next decade, regardless of their stage [5].

In Peru, up to 59.09% of people with type 2 diabetes are affected by CKD. We also know that it is the main cause of end-stage kidney disease and that it requires dialysis. The national rate of Peruvian prevalence per million population for renal replacement treatment is 335.3, with hemodialysis, peritoneal dialysis, and functional kidney transplant rates of 230.7, 39.1 and 65.5, respectively. Diabetes represents 40% of haemodialysis patients in a public hospital [6]. This requires an adequate capacity to evaluate and monitor the evolution of CKD in this specific population.

The use of glomerular filtration rate (GFR) is undoubtedly the most commonly used tool for detecting, managing and predicting the prognosis of CKD in patients with metabolic diseases such as type 2 diabetes mellitus [7]; however, some studies show that the methods for its estimation such as the formula MDRD (Modification of Diet in Renal Disease) or the formula of the CKD-EPI (Chronic Kidney Disease Epidemiology Collaboration) may be inadequate in diabetic patients due to an underestimation of these values [8,9].

In addition, other studies suggest that the values of Glycosylated Hemoglobin (HbA1c), an essential test for glycemic monitoring and control in diabetic patients, may be related to the variation in their GFR [10, 11], an inverse correlation between the increase in HbA1c and the decrease in GFR [12] was reported, even in the population of non-diabetic patients [13, 14]. A study assessing GFR predictive models using new parameters showed that the use of HbA1c has a high percentage of accuracy for predicting GFR [15].

The use of HbA1c to predict the variation of GFR would be very useful, given the need to find new alternatives for estimating GFR in diabetic patients without the need to increase costs, since these tests are part of the monitoring and control of all diabetic patients.

The aim of this study was to evaluate a predictive model for estimating GFR variability in diabetic patients based on HbA1c variability through a multiple linear regression analysis, incorporating into the analysis other variables that may influence GFR variability such as age, sex, blood pressure, BMI and routine laboratory tests.

## MATERIAL AND METHODS

The study is based on a cohort of patients with prospective follow-up belonging to the Adult Surveillance Program with Non-Transmission Chronic Disease, specifically, those who belong to the Renal Health Surveillance Program (VISARE) of the Hospital-I Pacasmayo of Essalud, La Libertad - Peru, a program that is implemented nationally for patients who are treated in Essalud.

This cohort has regular quarterly controls for the follow-up of patients with a diagnosis of Diabetes Mellitus type 2 with or without CKD, having a medical evaluation every 3 months, and monthly checks by the nursing staff for taking AP, BMI and programming of laboratory tests already stipulated as part of the program of Renal Health of the diabetic patient by Essalud, which include monthly evaluation of fasting glycemia, half-yearly evaluation of HbA1c, annual evaluation of lipid profile, hemoglobin, microalbuminurea, urine creatinine and serum creatinine.

The use of data was authorized by the Hospital I Pacasmayo to use data from the VISARE program and to have access to patients’ medical records. Information was subsequently collected from the hospital database for the period from 1 January 2014 to 31 December 2015.

We included patients who presented continuity in their medical controls for this program. The total number of patients identified according to the database obtained was submitted to the study selection criteria.

### Selection criteria

Inclusion criteria: Patients over 18 years of age with confirmed diagnosis of Diabetes Mellitus type 2 and belonging to the Renal Health surveillance program with at least an annual control in said program; patients who record age data, sex, laboratory tests at the time of periodic control in the program, including: serum creatinine, urine creatinine, microalbuminuria, hemoglobin, basal glycemia and HbA1c; diastolic blood pressure (DBP), systolic blood pressure (SBP), body mass index (BMI), GFR, use of nephroprotective drugs and comorbidities. Exclusion criteria: Patients with missing or poorly recorded data in any of the annual controls considered for the study.

After screening patients according to the selection criteria, information was collected from medical records. This information was encoded for analysis, which was executed using Stata 15.0 statistical software (StataCorp, College Station, TX, USA), performing a multiple linear regression that included the factors previously mentioned as possible predictors of GFR variability.

The result variable of the linear model was the variation of the TFG in the period of 1 year, which was calculated according to the values reported from the results for the years 2014 and 2015. Although many patients had quarterly controls, annual GFR variability was taken into consideration to better assess the plausibility of the factors included in this study, since evaluating short periods of these factors would have less influence on the variation of the TFG.

The variables blood pressure and basal glycemia were defined as the average of at least 3 measurements in consecutive months prior to the control by the program, due to their greater variability by various factors.

### Statistical analysis

The descriptive analysis was performed for each factor studied by calculating proportions for the categorical variables, as well as the mean, standard deviation and median for the numerical variables; additionally, the p value was reported to evaluate the difference between groups in the baseline analysis.

A Pearson correlation analysis was performed for variables of normal distribution and the Spearman test for variables without normal distribution. The analysis to determine the distribution of normality of the variables was performed using the Shapiro-Wilk test.

The inclusion of the factors in the multiple linear regression model was performed after evaluating the correlation between each factor studied and the variation of the GFR, the model was adjusted with the factors that showed to have a significant correlation.

### Ethical aspects

The research did not involve the direct participation of human beings (biological samples or interventions), it was worked with a hospital database authorized for use by the management of the institution. The handling of the information collected by the investigators was confidential and no personal data of any kind will be disclosed.

## RESULTS

We identified 234 patients, of whom only 122 diabetic patients met the selection criteria and were included in the multiple linear regression analysis to evaluate the predictive capacity of HbA1c, serum creatinine, and age. Of the study participants, the majority were male (55.7%) with no significant differences with respect to females (p= 0.20); the mean age was higher for men (67.4 years, C.I.: 65.2 - 69.6) than for women, there were significant differences between the two genders (p = 0.006). Of all patients, 63 (51.6%) had associated comorbidities, mainly hypertension (HT), with no difference between groups (p= 0.72); 76 (62.3%) used a nephroprotective drug (ACE inhibitor, ARA inhibitor or both), significantly different from those who did not use nephroprotective drugs (p= 0.006).

Regarding the BMI variation, there were no differences between both sexes (p= 0.16), with the male sex showing a variation of 0.02 (I.C.: -0.31, 0.28) and the female sex a variation of 0.33 (I.C.: -0.67, -0.001) (Table 1).

**Table 1:** Baseline of the study cohort of diabetic patients.

|  |  |  |  | 95% C.I. | p-value |
| --- | --- | --- | --- | --- | --- |
| **Gender, n (%)** | M | 68 | (55.7) | ... | 0.20 |
|  | F | 54 | (44.3) | ... |  |
| **Age (years), mean (SD)** | M | 67.4 | (9.0) | 65.2 – 69.6 | **0.006** |
|  | F | 61.2 | (10.6) | 58.3 – 64.1 |  |
|  | Total | 64.6 | (1.3) | 62.8 – 66.5 |  |
| **Co-morbidities, n (%)** | No | 59 | 48.4 | ... | 0.72 |
|  | Si | 63 | 51.6 | ... |  |
| **Nephroprotection, n (%)** | No | 46 | 37.7 | ... | **0.006** |
|  | Si | 76 | 62.3 | ... |  |
| **BMI change, mean** | M | -0.02 |  | -0.31, - 0.28 | 0.16 |
|  | F | -0.33 |  | -0.67, -0.01 |  |
|  | Total | -0.16 |  | -0.38, 0.06 |  |

C.I.: Confidence interval; SD: Standard deviation; BMI: Body Mass Index

Variables with normal distribution included Hb variation, serum creatinine variation, and HbA1c variation.

The correlation analysis showed that only HbA1c variation, serum creatinine variation, and microalbuminuria variation showed significant correlation when evaluated for GFR variability (Table 2).

**Table 2:** Correlation of the factors analyzed with respect to the variation of the GFR.

|  | Correlation coefficient | p-value |
| --- | --- | --- |
| Age | -0.1363 | 0.134 |
| Triglyceride variation | 0.0318 | 0.769 |
| Cholesterol variation | 0.0327 | 0.783 |
| LDL variation | 0.1507 | 0.276 |
| HDL variation | 0.0772 | 0.554 |
| Variation of serum creatinine | -0.8842 | **0.000** |
| Creatinuria Variation | -0.1331 | 0.144 |
| Variation of Micro albuminuria | -0.1973 | **0.029** |
| Variation of Alb/Cr rate | -0.1419 | 0.119 |
| Hemoglobina Variation | -0.0831 | 0.381 |
| BMI Variation | 0.1707 | 0.060 |
| Variation of Basal Glycemia | -0.0581 | 0.525 |
| Variation of SBP | 0.0448 | 0.624 |
| Variation of DBP | 0.0830 | 0.363 |
| Variation of HbA1c | -0.3059 | **0.000** |

LDL: Low density lipoprotein; HDL: High density lipoprotein; Alb/Cr : Albumin/Creatinine; BMI: Body Mass Index, SBP: systolic blood pressure; DBP: Systolic blood pressure, HbA1c: Glycated hemoglobin

**Table 3:** Multiple linear regression predictive model for GFR variation

|  | Model<br>coefficient | Standard<br>error | p-value of<br>the model | I.C. 95% |
| --- | --- | --- | --- | --- |
| HbA1c variation | -1.94 | 0.70 | 0.006 | -3.33 , -0.56 |
| Creatinine variation | -108.81 | 5.41 | 0.000 | -119.5 , -98.09 |
| Age | -0.04 | 0.02 | 0.009 | -0.07 , -0.01 |
| Adjusted R2: 0.8003, p < 0.0001. HbA1c: Glycated hemoglobin |  |  |  |  |

### Multivariate Analysis

the final multiple linear regression obtained after selecting the best adjusted model for predicting GFR variability was that which included HbA1c variation, serum creatinine variation, and age (Figures 1, 2, 3). The coefficients for the model were: -1.94 for HbA1c variation (p= 0.006), -108.81 for serum creatinine variation (p < 0.0001) and -0.04 for age (p= 0.009). The final model was very significant (p < 0.0001) with a variation of the HbA1 variance that explains 80% of the GFR (adjusted R2 = 0.80). The determination of the best model led to the elimination of the constant from the final formula as not being significant to the model.

**Figure 1:**
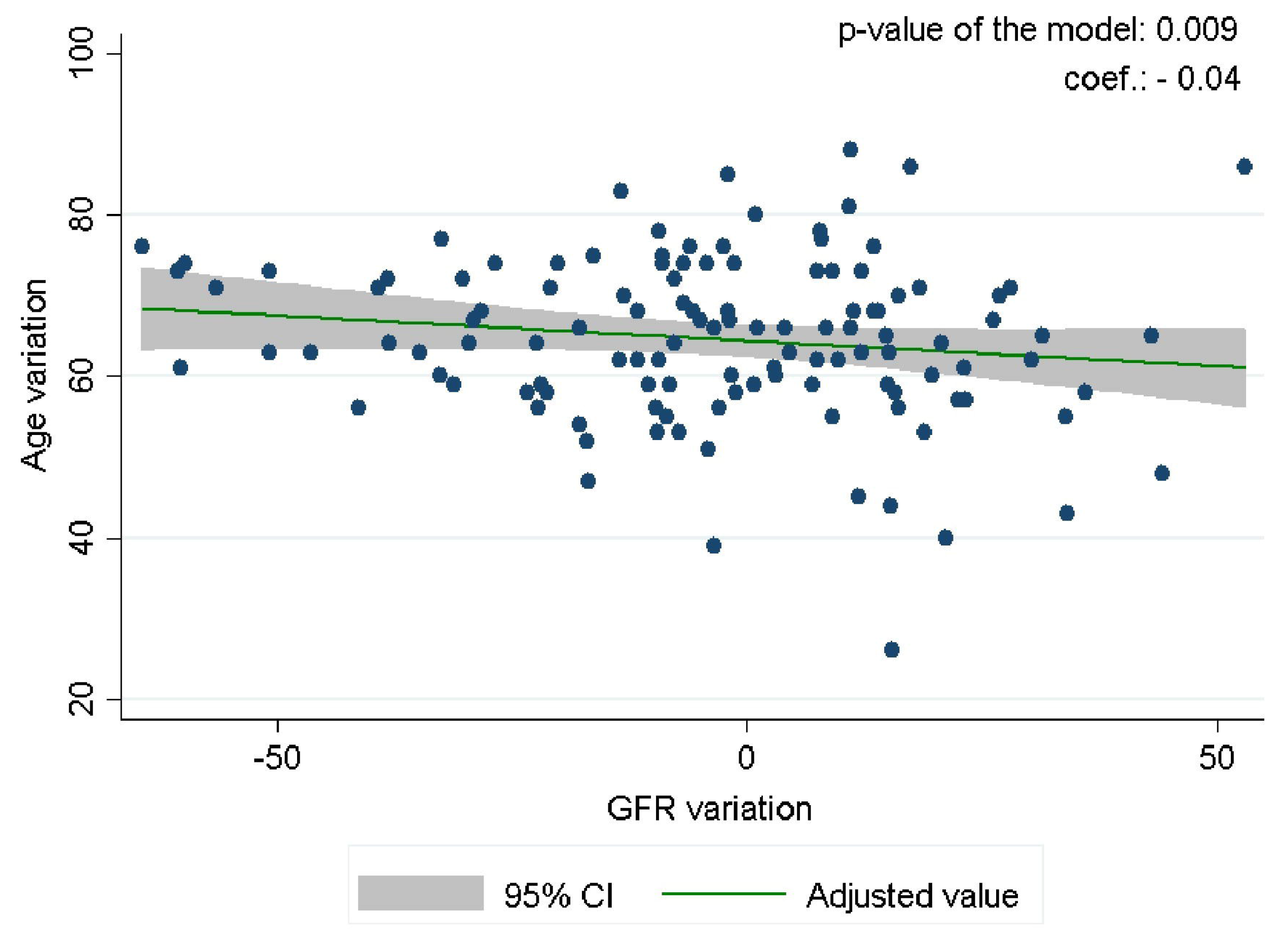
Linear prediction of GFR variation according to HbA1c variation

**Figure 2:**
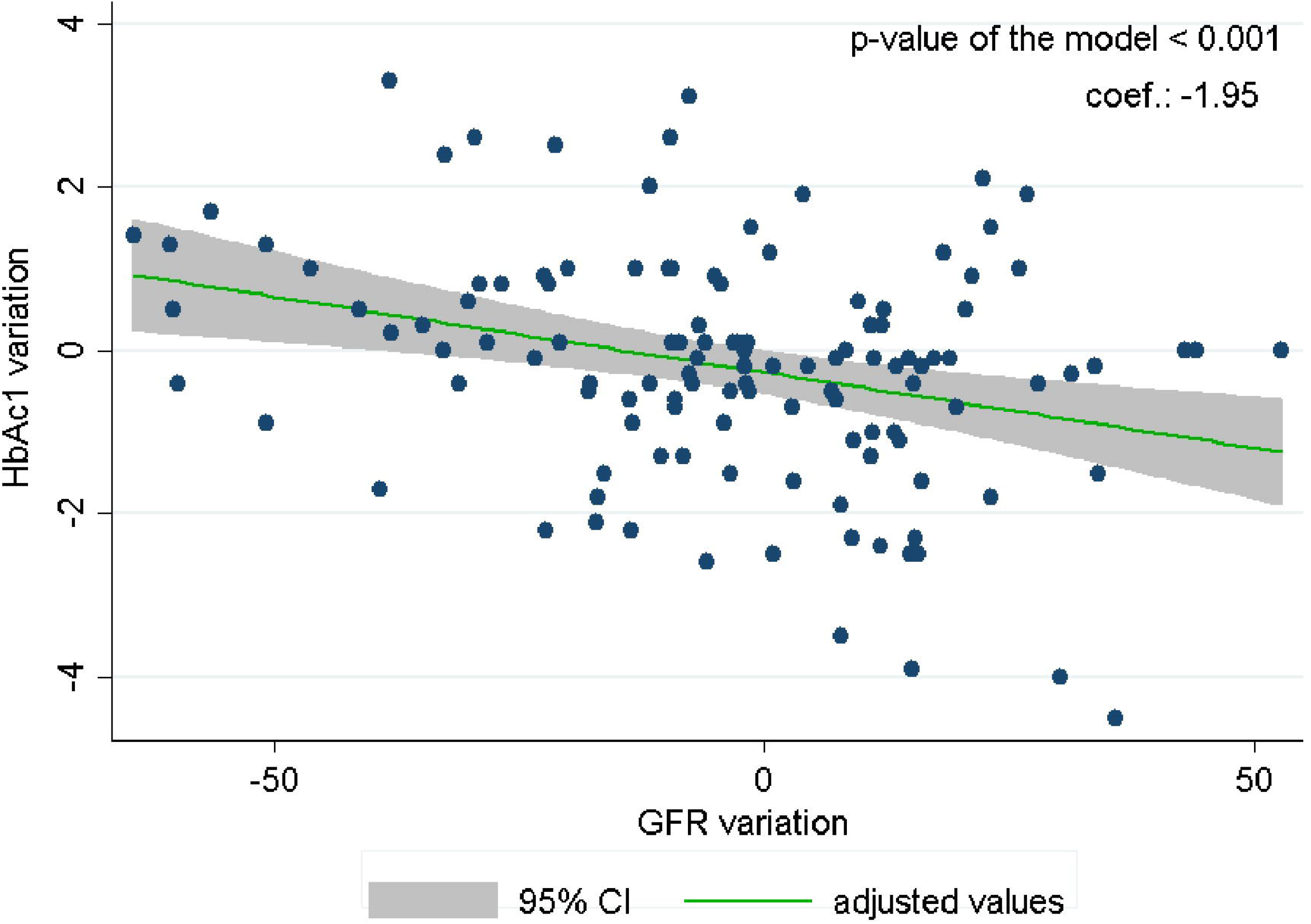
Linear prediction of GFR variation according to Creatinine variation

**Figure 3:**
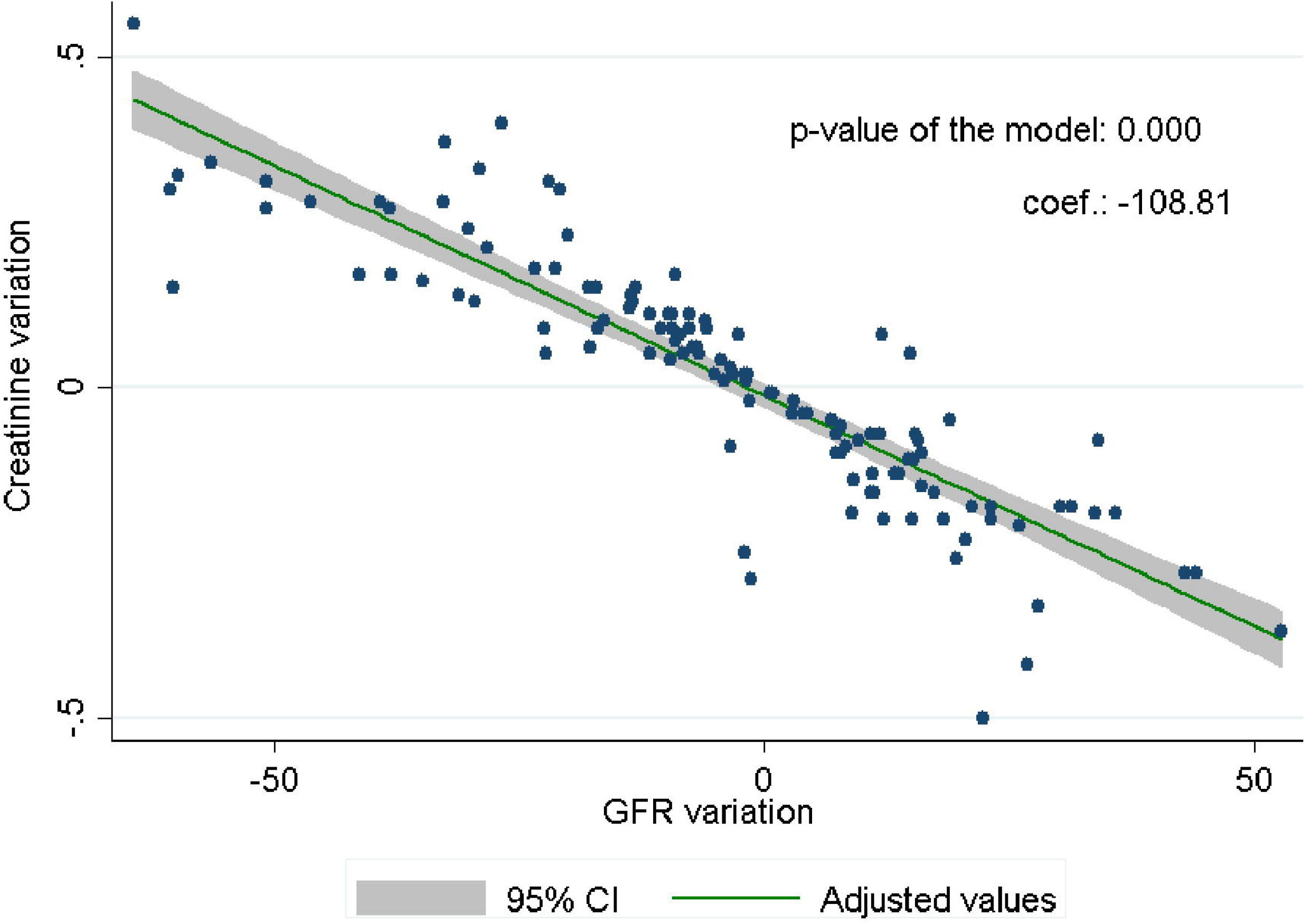
Linear prediction of GFR variation by age.

## DISCUSSION

The prevalence of diabetes is increasing, for 2014 it was estimated that there are 387 million cases worldwide [16], compared to previous decades [17]. There are no reliable data on the prevalence of diabetes in Peru, the few published reports show important percentages [18, 19]; of these, the percentage of patients with CKD is also important [6]. Diabetes in developed and some developing countries has become the leading cause of CKD. Therefore, it is necessary to accurately evaluate the GFR variation in diabetic patients, whether or not they have CKD.

Our analysis showed that HbA1c variation, serum creatinine variation, and age significantly predict GFR variation in diabetic patients regardless of their CKD condition. In research by Tsuda et al, a new equation was developed that includes HbA1c as a new variable. However, the sample size was very small (40 cases) [20]. The number of patients in our study was much higher, and the performance according to our study was very good.

The multiple regression model included an adequate number of patients, although the sample corresponded to a single center. We must also clarify that the variables, creatinine variability and age, are variables that show collinearity with the formula of Cockcroft - Gault and include them in the model was obviously necessary.

The results showing an explanatory level of 80% for the final model are quite satisfactory for this specific population, the model that proved to be very significant discusses the need to improve prognostic tools for CKD in these patients; such improvements should take into account the underlying disease and therefore try to use tests that are already part of the periodic control of the disease such as HbA1c.

It is important to note the limitations of our study. One of them is having performed the analysis on subjects that were restricted to a single center, under a renal control program. It is also important to mention that the study cohort was not completely uniform for age and nephroproprotection variables, there being a statistical difference when analyzed with respect to the sex of the patient, The average age in male patients is clearly higher than in the female sex, in the same way the proportion of patients who received some nephroproprotective drug is almost double that of those who do not receive any drug. These differences in baseline may have influenced the final model obtained since the relationship of GFR variation with age is known [21, 22], as well as the obvious effect of neproprotective drugs (ACE inhibitors and ARAs) on the evolution of GFR. Despite this, a very significant predictor model was achieved and it is necessary to emphasize that studies such as these are necessary in our environment, since many of the GFR estimation formulas are based on foreign populations with very different realities in terms of the parameters commonly used and the same idiosyncrasy of each individual.

## CONCLUSIONS

The optimal study model for predicting GFR variation in diabetic patients with or without CKD included HbA1c, serum creatinine variation, and age, with a high percentage of predictability, however it is important to note that the HbA1c correlation level was low and there were factors that could have affected the results in the study population, such as the differentiated use of nephroproprotective drugs by much of the study cohort and the age difference by sex. New research studies are required that consider a much larger sample and standardize criteria to eliminate confounding factors that could alter the predictive capacity of HbA1c for prognosis of GFR variation. It is possible to further improve the predictive capacity of HbA1c by improving the analysis methodology.

## Data Availability

All data produced in the present study are available upon reasonable request to the authors

## REFERENCES

1. Thomas MC, Cooper ME, Zimmet P. Changing epidemiology of type 2 diabetes mellitus and associated chronic kidney disease. Nat Rev Nephrol. February 2016;12(2):73–81.

2. Metsärinne K, Bröijersen A, Kantola I, Niskanen L, Rissanen A, Appelroth T, et al. High prevalence of chronic kidney disease in Finnish patients with type 2 diabetes treated in primary care. Prim Care Diabetes. February 2015;9(1):31–8.

3. Prasannakumar M, Rajput R, Seshadri K, Talwalkar P, Agarwal P, Gokulnath G, et al. An observational, cross-sectional study to assess the prevalence of chronic kidney disease in type 2 diabetes patients in India (START - India). Indian J Endocrinol Metab. August 2015;19(4):520–3.

4. Janmohamed MN, Kalluvya SE, Mueller A, Kabangila R, Smart LR, Downs JA, et al. Prevalence of chronic kidney disease in diabetic adult out-patients in Tanzania. BMC Nephrol. 2013;14:183.

5. Kainz A, Hronsky M, Stel VS, Jager KJ, Geroldinger A, Dunkler D, et al. Prediction of prevalence of chronic kidney disease in diabetic patients in countries of the European Union up to 2025. Nephrol Dial Transplant Off Publ Eur Dial Transpl Assoc - Eur Ren Assoc. August 2015;30 Suppl 4:iv113–118.

6. Villena JE. Diabetes Mellitus in Peru. Ann Glob Health. December 2015;81(6):765–75.

7. Levey AS, Inker LA, Coresh J. GFR estimation: from physiology to public health. Am J Kidney Dis Off J Natl Kidney Found. May 2014;63(5):820–34.

8. Silveiro SP, Araújo GN, Ferreira MN, Souza FDS, Yamaguchi HM, Camargo EG. Chronic Kidney Disease Epidemiology Collaboration (CKD-EPI) equation pronouncedly underestimates glomerular filtration rate in type 2 diabetes. Diabetes Care. November 2011;34(11):2353–5.

9. Camargo EG, Soares AA, Detanico AB, Weinert LS, Veronese FV, Gomes EC, et al. The Chronic Kidney Disease Epidemiology Collaboration (CKD-EPI) equation is less accurate in patients with Type 2 diabetes when compared with healthy individuals. Diabet Med J Br Diabet Assoc. January 2011;28(1):90–5.

10. Lee C-L, Li T-C, Lin S-Y, Wang J-S, Lee I-T, Tseng L-N, et al. Dynamic and dual effects of glycated hemoglobin on estimated glomerular filtration rate in type 2 diabetic outpatients. Am J Nephrol. 2013;38(1):19–26.

11. Solini A, Manca ML, Penno G, Pugliese G, Cobb JE, Ferrannini E. Prediction of Declining Renal Function and Albuminuria in Patients With Type 2 Diabetes by Metabolomics. J Clin Endocrinol Metab. February 2016;101(2):696–704.

12. Joly D, Choukroun G, Combe C, Dussol B, Fauvel J-P, Halimi J-M, et al. Glycemic control according to glomerular filtration rate in patients with type 2 diabetes and overt nephropathy: a prospective observational study. Diabetes Res Clin Pract. April 2015;108(1):120–7.

13. Pallayova M, Mohammed A, Langman G, Taheri S, Dasgupta I. Predicting non-diabetic renal disease in type 2 diabetic adults: The value of glycated hemoglobin. J Diabetes Complications. July 2015;29(5):718–23.

14. Trivin C, Metzger M, Haymann J-P, Boffa J-J, Flamant M, Vrtovsnik F, et al. Glycated Hemoglobin Level and Mortality in a Nondiabetic Population with CKD. Clin J Am Soc Nephrol CJASN. June 2015;10(6):957–64.

15. Chen J, Tang H, Huang H, et al. Development and validation of new glomerular filtration rate predicting models for Chinese patients with type 2 diabetes. J Transl Med. 2015;13:317. Published 2015 Sep 28. doi:10.1186/s12967-015-0674-y

16. IDF diabetes atlas. 10^th^ Edition. Available in: http://www.diabetesatlas.org/

17. Danaei G, Finucane MM, Lu Y, Singh GM, Cowan MJ, Paciorek CJ, et al. National, regional, and global trends in fasting plasma glucose and diabetes prevalence since 1980: systematic analysis of health examination surveys and epidemiological studies with 370 country-years and 2·7 million participants. Lancet Lond Engl. July 2011;378(9785):31–40.

18. Ramos W, López T, Revilla L, More L, Huamaní M, Pozo M.. Results of the epidemiological surveillance of diabetes mellitus in hospitals in Peru, 2012. Rev Peru Med Exp Salud Pública. 2014;31(1):9–15.

19. Goldhaber-Fiebert JD, Jeon CY, Cohen T, Murray MB. Diabetes mellitus and tuberculosis in countries with high tuberculosis burdens: individual risks and social determinants. Int J Epidemiol. April 2011;40(2):417–28.

20. Tsuda A, Ishimura E, Ohno Y, Ichii M, Nakatani S, Machida Y, et al. Poor glycemic control is a major factor in the overestimation of glomerular filtration rate in diabetic patients. Diabetes Care. 2014;37(3):596–603.

21. Cohen E, Nardi Y, Krause I, Goldberg E, Milo G, Garty M, et al. A longitudinal assessment of the natural rate of decline in renal function with age. J Nephrol. December 2014;27(6):635–41.

22. Wang X, Vrtiska TJ, Avula RT, Walters LR, Chakkera HA, Kremers WK, et al. Age, kidney function, and risk factors associate differently with cortical and medullary volumes of the kidney. Kidney Int. March 2014;85(3):677–85.

